# Geoepidemiology of COVID-19 hospitalisations and severity: impact of social deprivation and remote areas, in the south-eastern Region, France

**DOI:** 10.1101/2025.11.17.25340377

**Authors:** Pierre Garneret, Guillaume Gaubert, Steve Nauleau, Florian Franke, Stanislas Rebaudet, Emilie Mosnier, Luka Canton, Pierre Schalkwijk, Pascal Chaud, Sabira Smaïli, Stéphanie Vandentorren, Michael Huart, Marc-Karim Bendiane, Jordi Landier, Jean Gaudart

## Abstract

**Background:** Since the onset of COVID 19 pandemic, multiple individual-level factors, including socioeconomic determinants have been associated with infection risk and disease severity. However, public health policies implemented at national level did not consider social determinants at territorial and community levels. In collaboration with the Regional Health Agency of Provence-Alpes-Côte d’Azur (PACA) in France, we analysed COVID-19 hospitalisations together with incidence and testing data, at a fine geographical scale, in order to contextualise the epidemic, and assess the impact of area-level socioeconomic and demographic characteristics on disease severity.

**Methods:** We conducted a fine scale ecological study of COVID-19 hospitalisation rates in the PACA region, during the second and third epidemic waves (September 2020-June 2021). French census areas (IRIS), the smallest spatial units available for socioeconomic and population-based analysis, were classified into six socio-demographic profiles. We characterized COVID-19 with indicators of incidence and severity relative to total population (incidence and hospitalisation rates) and proportional to tests or cases (proportion of positive tests and proportion of hospitalised cases). Associations between these profiles and COVID-19 indicators were assessed using generalised additive models, adjusting for testing rates, healthcare access, retirement home presence and population age structure. Spatial autocorrelation between areas was accounted for in the models.

**Results:** The most socially deprived IRIS had the highest COVID-19 incidence and hospitalisation rates both in conventional settings and in intensive care units (ICU). Decreasing social deprivation was associated with a gradient of decreasing incidence and hospitalisation rates. Complementary models examining proportion of hospitalisation among confirmed cases indicated that excess hospitalisation in very socially deprived area reflected both higher incidence and greater severity of the disease. IRIS profiles corresponding to remote, rural areas displayed an isolated increase in conventional hospitalisation ratios without a corresponding rise in ICU hospitalisation ratios.

**Conclusions:** Socioeconomic deprivation was strongly associated with both higher infection spread and greater severity of COVID-19 at the territorial level, underscoring the need to prioritize prevention efforts in socially deprived areas to mitigate future health crisis. Remote areas also exhibited higher conventional hospitalisation rates, possibly reflecting clinical decisions influenced by remoteness rather than increased disease severity.

**Graphical abstract:** 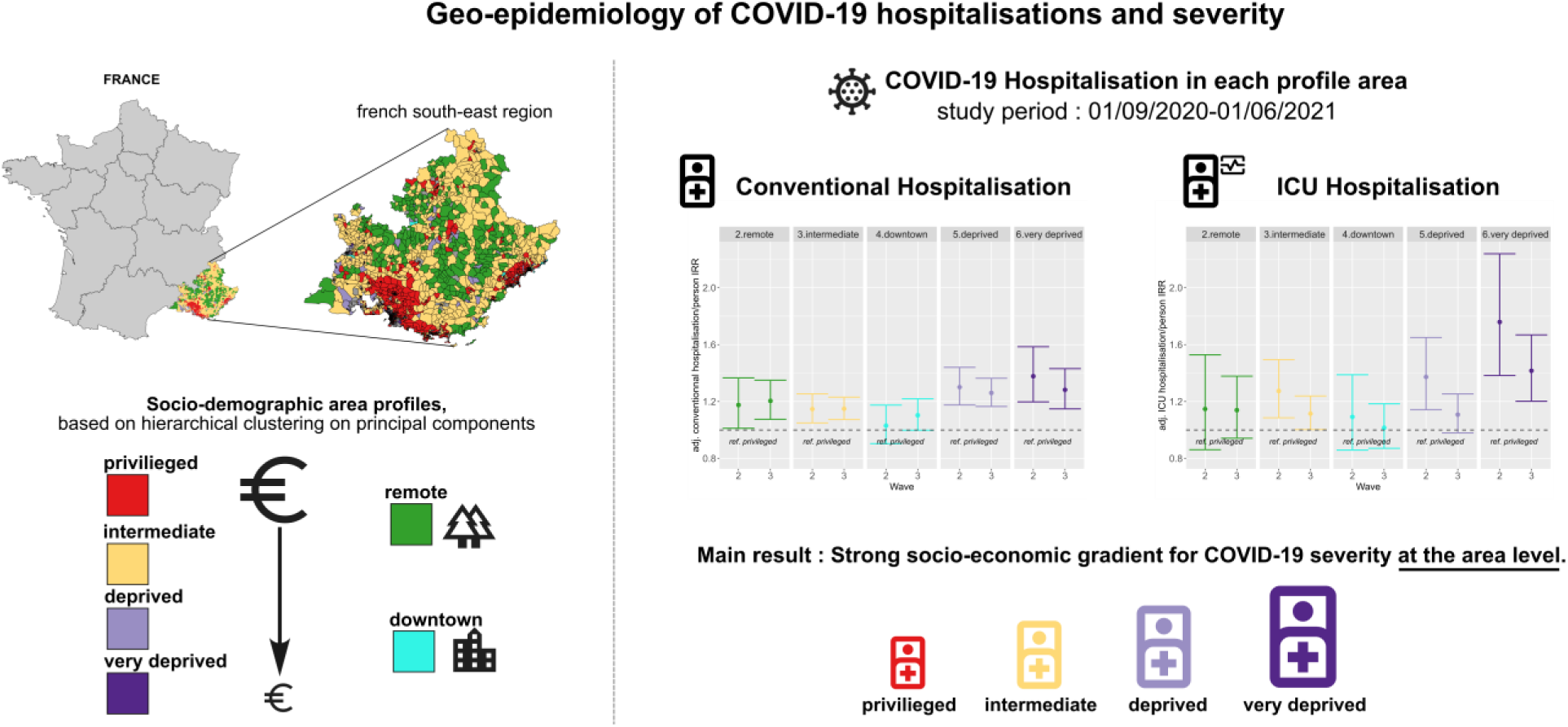

## Introduction

Since its emergence, the spread and severity of COVID-19 have been associated with a wide range of contextual factors, including population age structure (1), healthcare system capacity and accessibility (2), climatic conditions (3), and the timing and stringency of non-pharmaceutical interventions (4,5). Among these, socioeconomic inequalities have consistently emerged as a key determinant across several countries (6–8).

These multidimensional inequalities –encompassing social class, gender, ethnicity and education (9,10)—have been shown to influence both COVID-19 transmission dynamics and disease severity at the individual level (11–13). Identifying the territories where such inequalities translate into greater disease burden requires fine-scale spatial analysis. Ecological approaches are particularly valuable in this regard, as they allow quantification of the relationship between deprivation and health outcomes at the local scale, while also delineating areas where targeted interventions should be prioritised (14–17). Furthermore, fine-scale characterisation captures regional and demographic specificities, which may not be fully represented by nationally standardised socioeconomic indices (18).

In France, fine-scale dynamics of COVID-19 testing and incidence rates have been previously investigated (18–20). These studies showed that, even within a healthcare system designed to ensure universal coverage, the socioeconomic profile of a given area strongly influenced testing behaviour and viral spread. Limitations in health care accessibility, particularly in remote and rural areas, were also found to affect testing dynamics. To date, however, COVID-19 severity – measured through conventional or intensive care unit (ICU) hospitalisations – has been far less studied at this local level.

The Provence-Alpes-Côte d’Azur (PACA) region, in south-eastern France combines densely populated coastal cities with remote rural and mountainous areas. Urban areas also display pronounced socioeconomic inequalities between the most affluent and the most deprived neighbourhoods. Before the widespread implementation of vaccination campaigns, the region was heavily affected by the COVID-19 pandemics, and its regional databases provides detailed information on hospitalisations (21).

In the present study, we focused on the second and third waves of COVID-19 in PACA (September 2020 to June 2021), aiming to investigate the association between socioeconomic and demographic profiles and hospitalisation dynamics, at a fine geographical scale.

## Material and methods

### Study design

This ecological population-based study aimed to assess the impact of socio-demographic characteristics on COVID-19 incidence and hospitalisation at a fine geographical scale. The PACA region is located in south-eastern France with a population of 5 million inhabitants, of which 88% are concentrated in urban areas along the Mediterranean coast (22). The *Regrouped Islet for statistical information* (IRIS) correspond to census blocks of approximately 1,000 to 5,000 inhabitants and represent the finest statistical unit available in France. Analyses were performed at the IRIS level. In addition to social deprivation, age structure and healthcare services provision were examined as covariates.

### Study period

We analysed the period between 2020-09-01 and 2021-06-01 corresponding to the second and third epidemic waves in France. Earlier data were excluded due to limited standardization in hospital reporting, whereas later data were not considered because the rollout of vaccination campaigns could confound interpretation of the results. The cut-off date between the two waves was set at December 15, 2020 which marked the end of the nationwide lockdown initiated in November, coincided with the start of school holidays family gathering during the year end period, and was characterised by a temporary decline in COVID-19 incidence.

### IRIS selection criteria

According to INSEE, IRIS are classified into three categories: “residential”, “employment/industrial”, and “special purpose”. We excluded “employment/industrial” IRIS which mainly represent the working rather than the resident population as well as all “special purpose” and “residential” IRIS with fewer than 30 inhabitants, in accordance with French data protection regulations to avoid any risk of individual identification. In addition, IRIS with no hospitalisation data were excluded from the analysis.

### Outcomes

All COVID-19 cases detected in the population were extracted from the national information system implemented on May 13, 2020 by Sante publique France (the French National Agency of Public Health), called SI-DEP (“Système d’Information de Dépistage Populationnel”). SI-DEP is a secure platform that records all SARS-CoV-2 RT-PCR and antigen test results from laboratories, hospitals, pharmacies, nurses, and physicians across France, including information on sex, age, and residential address.

Hospital admissions of COVID-19 diagnosed patients were reported by hospital services to Santé publique France, through another national information system called SI-VIC (“Système d’Information pour le suivi des VICtimes”). Two types of hospitalisations were considered: “conventional hospitalisation” (i.e., in non-critical settings) and “intensive care units (ICU) hospitalisation” (including, in this study, resuscitation, intensive care and continuing care units). The indications for hospitalisation were based on the observation of severity criteria such as polypnea or desaturation, and critical care was indicated when permanent monitoring was necessary (23–25).

Data aggregation was performed by the regional health authority, (“Agence Régionale de Santé, Provence Alpes Cote d’Azur”, ARS-PACA) for the cases and hospital admissions, which linked residential address of COVID-19 cases or admitted patients to the corresponding IRIS after geocoding. The level of temporal aggregation was the wave as previously defined, according to the rolling week of screening for cases and tests, and the day of admission for hospital data.

In order to analyse the occurrence of hospitalisations, we had to first characterise viral spread at a fine temporal scale, and we considered three complementary indicators: (i) the daily COVID-19 testing rate, (ii) the daily COVID-19 incidence rate and (iii) the daily COVID-19 positivity rate. Together, these measures provided a comprehensive picture of transmission dynamics and testing behaviours.

Daily hospitalisation-to-population incidence rate were calculated separately for conventional and ICU admission. This rate was defined as the ratio of hospitalisation (conventional or ICU) to the number of inhabitants in the corresponding IRIS, on a given day. To account for reporting fluctuation linked to weekends, hospitalisation incidence curves were smoothed using a 7-day moving average.

For the model-based approach, outcomes were analysed at the level of each epidemic wave. We considered (i) the COVID-19 incidence rate and (ii) the hospitalisation-to-population rates (in conventional or ICU settings) for the main analysis. Complementary analysis considered (iii) the proportion of positive tests among all tests performed, and (iv) the proportion of hospitalised cases (in conventional or ICU settings) among all confirmed cases (Table 1).

**Table 1:** Definition of the COVID-19 indicators used for the statistical modelling.

|  | Population-based incidence | Proportions |
| --- | --- | --- |
| **Case load** | COVID-19 incidence rate<br>$\frac{\text{Number of confirmed cases}}{\text{Exposed person. Time}}$ | Proportion of positive tests<br>$\frac{\text{Number of confirmed cases}}{\text{Total tests}}$ |
| **Conventional hospitalisation** | Conventional hospitalisation-to-population rate<br>$\frac{\text{Number of conventional hospitalisation}}{\text{Exposed person. time}}$ | Proportion of hospitalised cases in conventional settings<br>$\frac{\text{Number of conventional hospitalisation}}{\text{Total confirmed cases}}$ |
| **ICU hospitalisation** | ICU hospitalisation-to-population rate<br>$\frac{\text{Number of ICU hospitalisation}}{\text{Exposed person. time}}$ | Proportion of hospitalised cases in ICU settings<br>$\frac{\text{Number of ICU hospitalisation}}{\text{Total confirmed cases}}$ |
| **Location in the article** | Fig 3 and Tables S2,S3,S4 | Fig S5 and Tables S5,S6,S7 |

### Variables

Detailed data sources are detailed in Table S1. Population socio-demographic data were obtained from the national Institute of Economic Studies (INSEE) open data portal at the IRIS level (Regrouped Islets for Statistical Information) (26). The 2017 European deprivation Index (EDI) by IRIS was provided obtained by MapInMed upon request.

Social deprivation is defined in relative terms within a given population (27), and we relied on an unsupervised categorisation of IRIS into 6 socio-demographic profiles, as previously described by Landier et al. (18). Briefly, we constructed profiles using hierarchical ascendant classification based on principal component analysis coordinates (HCPC). This unsupervised method, incorporated the following variables: population density, proportion of each socio-professional category, proportion of foreigners and immigrants, bachelor’s degree rate, unemployed rate, median income, European Deprivation Index (28), and proportion of overcrowded homes. It is designed to reduce issues related to dimensionality and collinearity (29). These six profiles are structured around two main components: deprivation level, and population density (see Fig. S1 and more details in (18)).

- The first profile, characterised by high median income and intermediate population density, located in urban and peri-urban areas, was defined as “privileged”. It clustered around Aix-en-Provence and along the coastal corridor between Marseille and Toulon, as well as between Cannes and Nice
- Three middle-income profiles were distinguished based on population density and spatial distribution: “remote” (rural areas, low density, relatively high proportion of agricultural workers, predominantly located in mountainous areas), “intermediate” (intermediate density, urban and peri-urban areas) and “downtown” (central districts of major cities, high density).
- A high-density urban profile with lower median income and a large proportion of blue-collar workers was defined as “deprived”.
- Finally, the “very deprived” profile corresponded to urban areas exhibiting highest densities and highest European deprivation index (EDI), typically characterised by large-scale social housing projects. “Deprived” and “very deprived” IRIS were concentrated in the northern districts of Marseille and in specific neighbourhoods of Nice and Toulon.

In order to reflect the demographic composition of local populations, we used a previously HCPC defined 4-class age-structure profile, based on the proportion of each age category within each IRIS (proportion of different age group [< 18; 18-39; 40-64; ≥ 65]). This classification identified four distinct profiles. The first three – “families”, “young adults”, and “elderly”—were characterised by higher proportions of individuals aged < 18 years, 18-39 years, and ≥ 65 years respectively. The fourth profile termed “balanced”, displayed a relatively even distribution across all age categories.

Healthcare service provision was characterised using four variables: (i) presence of a biological laboratory (common site for COVID-19 testing); (ii) number of frontline caregivers (including nurses, general practitioners, gynecologists, pediatricians, ophthalmologists, psychiatrists, dental surgeons, midwives, physiotherapists) (Fig S3 A); (iii) distance between IRIS centroid and nearest emergency department (in kilometres) (Fig S3 B); and (iv) localised potential access to a general practitioner (LPA) measured by the number of possible consultations per person and per year (30). This last variable was only available at the municipality-level.

The presence of a retirement home within the IRIS was also included as an adjustment variable to account for local concentration of older and more vulnerable populations.

### Statistical analysis

For the model-based approach, the main analysis considered the COVID-19 incidence rate and the hospitalisation-to-population rates, estimated separately for conventional and ICU settings. The hospitalisation-to-population model was expected to capture accurately severity heterogeneities between areas, but it could be confounded by differential viral circulation. To disentangle the respective contribution of incidence and disease severity, a complementary set of analyses was performed modelling the proportion of positive test and the proportion of hospitalised cases.

COVID-19 incidence rate and hospitalisation-to-population rates were mapped for each IRIS and epidemic wave. Spatial autocorrelation between IRIS was quantified using Moran’s I statistic, based on the queen’s contiguity criterion (31).

For each epidemic wave (second and third), associations between potential determinants (socio-demographic and age structure profiles, and health services provision) and COVID-19 indicators were estimated using Generalized Additive Mixed Models (GAMM). The “privileged” and “young adult” profiles were used as reference categories for the socio-demographic and age structure classifications, respectively. A negative binomial distribution (NegBin) was specified to account for overdispersion in hospital admission data. A random effect at the municipality level (RE[city]) was introduced, to account for the scale at which LPA was measured. Spatial correlation was further addressed by applying a Gaussian kriging smoother to the geographical coordinates of IRIS centroids (s[longitude, latitude]) (32).

The final models were specified as follows:

Incidence rate:

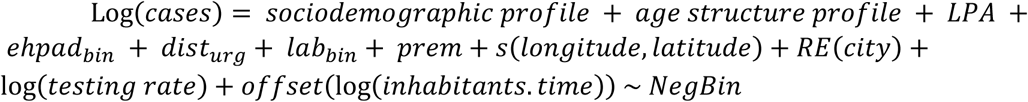

Conventional/ICU hospitalisation-to-population rate:

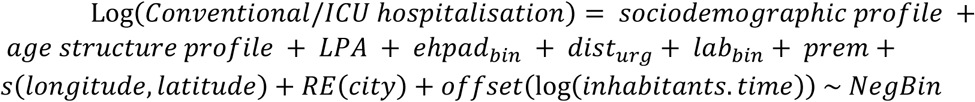

Proportion of positive test:

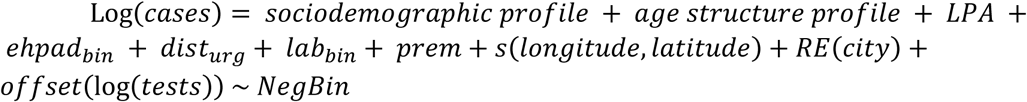

Proportion of cases hospitalised in conventional/ICU settings:

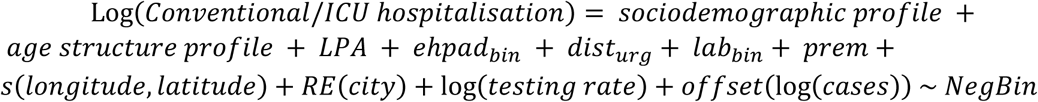

The spatial autocorrelation of the model residuals was assessed using Moran’s I statistic as a sensitivity analysis, in order to confirm the absence of residual spatial dependence.

All analyses and cartographic representations were performed with R software version 4.5.0 (R Development Core Team, R Foundation for Statistical Computing, Vienna, Austria) using in particular the following packages: {FactoMineR}(29), {mgcv}(33), and {spdep}(34).

All data produced in the present study are available upon reasonable request to the authors

## Results

### IRIS selection

The PACA region comprises 2,446 IRIS corresponding to 5.04 million inhabitants. A total of 139 IRIS were excluded due to their “employment/industrial” category or because they contained fewer than 30 inhabitants. In addition, 177 IRIS with no hospitalisation data were removed. The final analytical dataset included 2,130 IRIS, covering 4.92 million inhabitants (Fig S2).

### Time evolution of COVID-19 incidence and hospitalisations by socio-demographic profile

To provide a comprehensive view of COVID-19 dynamics and severity, we examined daily incidence rates, along with daily hospitalisation rates in conventional and ICU settings (Fig 1). We also present daily testing rates analysed previously (18) since testing is the source of information on cases and a potential source of bias in characterizing the dynamics.

**Figure 1:**
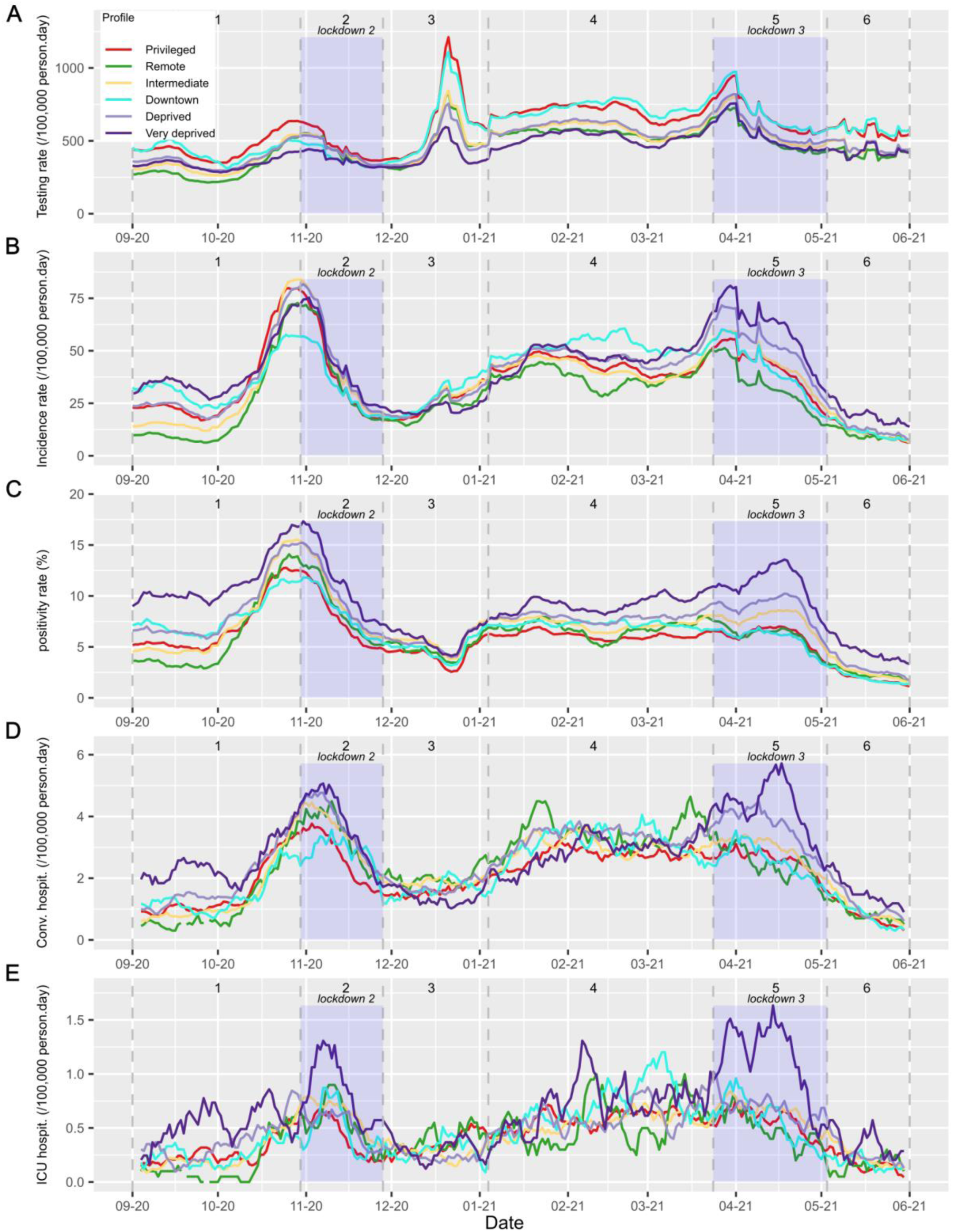
Evolution of daily COVID-19 (A) testing rate, (B) incidence rate, (C) positivity rate, (D) conventional hospitalisation rate and (E) ICU hospitalisation rate, by socio-demographic profile in PACA region, from September 2020 to June 2021. For finer temporal stratification and enhanced contextualization, data were represented across six pre-defined periods based on Landier et al. (18): (1) wave 2 rising (01/09/2020-29/10/2020), (2) Lockdown 2 (30/10/2020-27/11/2020), (3) Christmas lull (28/11/2020-03/01/2021), (4) wave 3 rising (04/01/2021-23/03/2021), (5) Lockdown 3(24/03/2021-02/05/2021), (6) wave 3 falling (03/05/2021-01/06/2021). This allowed for more detailed contextualisation description of COVID 19 indicators across the two epidemic waves.

Testing activity displayed three distinct peaks: two aligned with the incidence peaks and one during the Christmas period. Privileged and downtown IRIS consistently recorded the highest testing rates, while very deprived showed the lowest (Fig 1A).

Across all profile, incidence rate followed a two-wave pattern (Fig 1B). Remote IRIS displayed lower incidence throughout the study period. Wave 2 incidence peaks were more pronounced in privileged, intermediate and deprived IRIS, whereas wave 3 was driven by higher incidence in very deprived, deprived and downtown IRIS. Notably, during lockdown 3, very deprived and deprived maintained the highest rates, despite lower testing activity. Positivity rates, as a result, exhibited consistently higher values for very deprived and deprived IRIS, particularly during epidemic peaks corresponding to lockdown 2 and lockdown 3 (Fig 1C).

Conventional hospitalisation rates closely followed the kinetics of incidence and positivity rates with a short delay (Fig 1D). During the rise of the second wave and the implementation of the second lockdown, very deprived IRIS exhibited higher rates. In contrast, at the onset of the third wave, remote IRIS show the highest rates. During the third lockdown, hospitalisation rates in remote IRIS declined rapidly, whereas both very deprived and deprived IRIS maintained elevated rates, which continued to increase throughout the lockdown.

ICU hospitalisation rates were predominantly higher in very deprived IRIS, with elevated values observed during the rise of the second wave, the second lockdown, and the third lockdown (Fig 1E). By comparison, remote IRIS did not exhibit higher ICU admission rates

Overall, very deprived IRIS consistently showed higher positivity rates, as well as elevated conventional and ICU hospitalisation rates. These differences compared with privileged IRIS were more pronounced during the rise of the second wave, the second lockdown, and the third lockdown. Remote IRIS generally displayed lower incidence and positivity rates but exhibited higher conventional hospitalisation rates particularly during Christmas period and at the onset of the third wave. This pattern was not observed for ICU admission.

### Spatial distribution of COVID-19 incidence and hospitalisation rates

Incidence rates displayed distinct spatial patterns across the two waves (Fig 2A). During wave 2, higher incidence was observed in the western part of the region, whereas wave 3 predominantly affected the eastern coastal areas, particularly around Fréjus, Cannes and Nice. Marseille was strongly impacted during both waves. Spatial autocorrelation, as measured by Moran’s I, was significant for both waves, with values of 0.29 and 0.41, respectively (Table S8).

**Figure 2:**
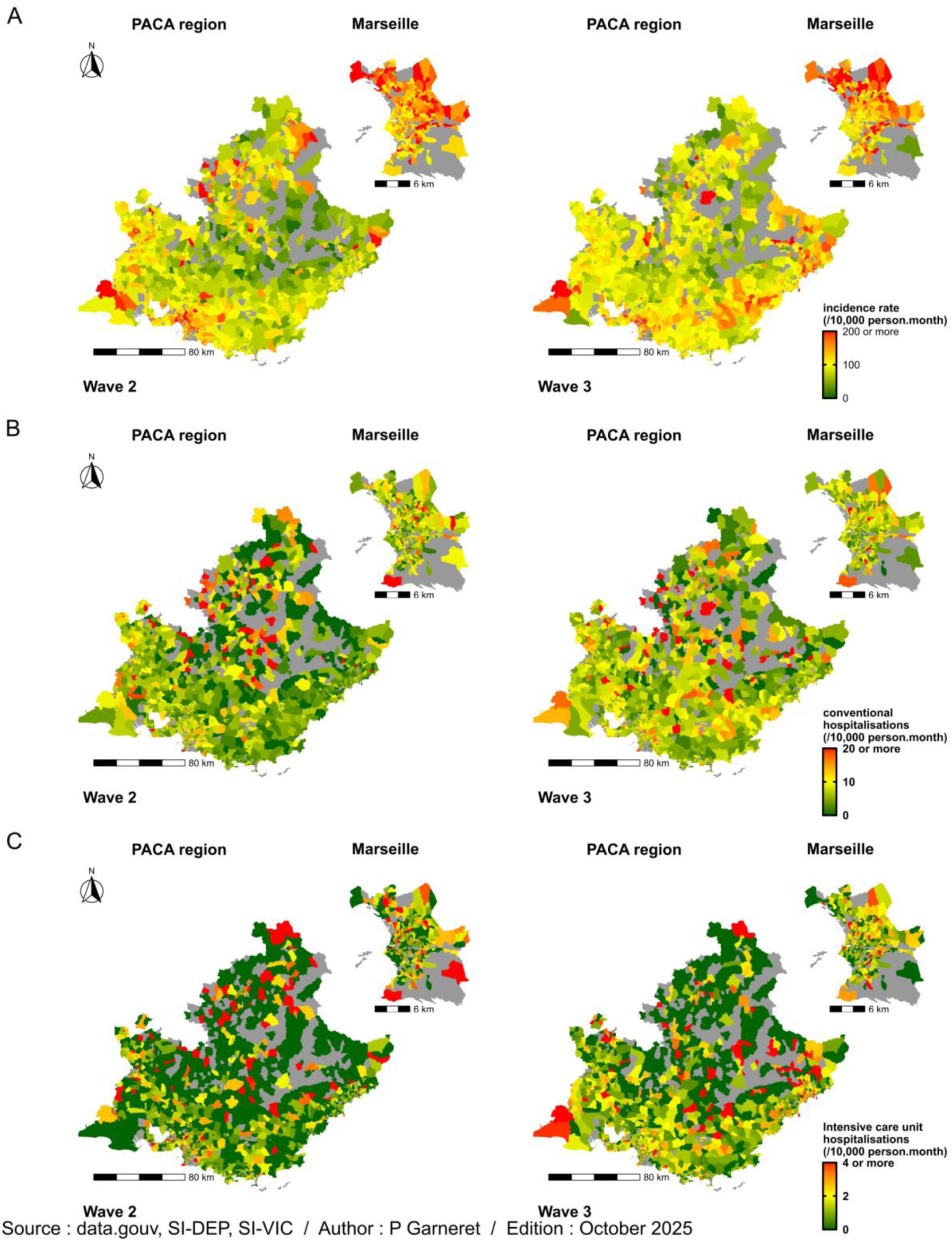
Spatial distribution of COVID-19 (A) incidence rate, as well as (B) conventional and (C) ICU hospitalisation rates in the PACA region, with a focus on Marseille, during epidemic waves 2 and 3. Grey areas correspond to IRIS unit with no hospitalisation data reported.

Positivity rates showed a more pronounced geographical distribution (Fig S4). A north-west to south-east gradient was evident in wave 2, with higher values in the western part of the region. This gradient was no longer apparent during wave 3. Within Marseille, positivity rates remained consistently higher in northern districts, corresponding to low resource neighbourhoods, across both waves.

Conventional hospitalisation rates were higher in the north-western IRIS during wave 2 and more evenly distributed during wave 3 (Fig 2B). At the city scale, no specific spatial clustering was observed in Marseille. Moran’s I indicated significant spatial autocorrelation for both waves (0.16 and 0.08 respectively; Table S8)

ICU hospitalisation rates did not show a clear geographical pattern, although a tendency towards higher values in coastal IRIS was observed during wave 3 (Fig 2C). Spatial autocorrelation was significant for this wave (Moran’s I = 0.10; Table S8)

### Factors associated with COVID-19 incidence and hospitalisations

After adjustment for spatial autocorrelation, testing rate, healthcare access variables and age structure, very deprived IRIS showed significantly higher COVID-19 incidence rate ratios (IRR) and hospitalisation-to-population rate ratio (HRR) in both conventional settings (1.38 [1.20-1.59]; 1.28 [1.15-1.43]) and ICU settings (1.76 [1.38-2.24]; 1.42 [1.20-1.67]) (Fig 3, Table S2).

**Figure 3:**
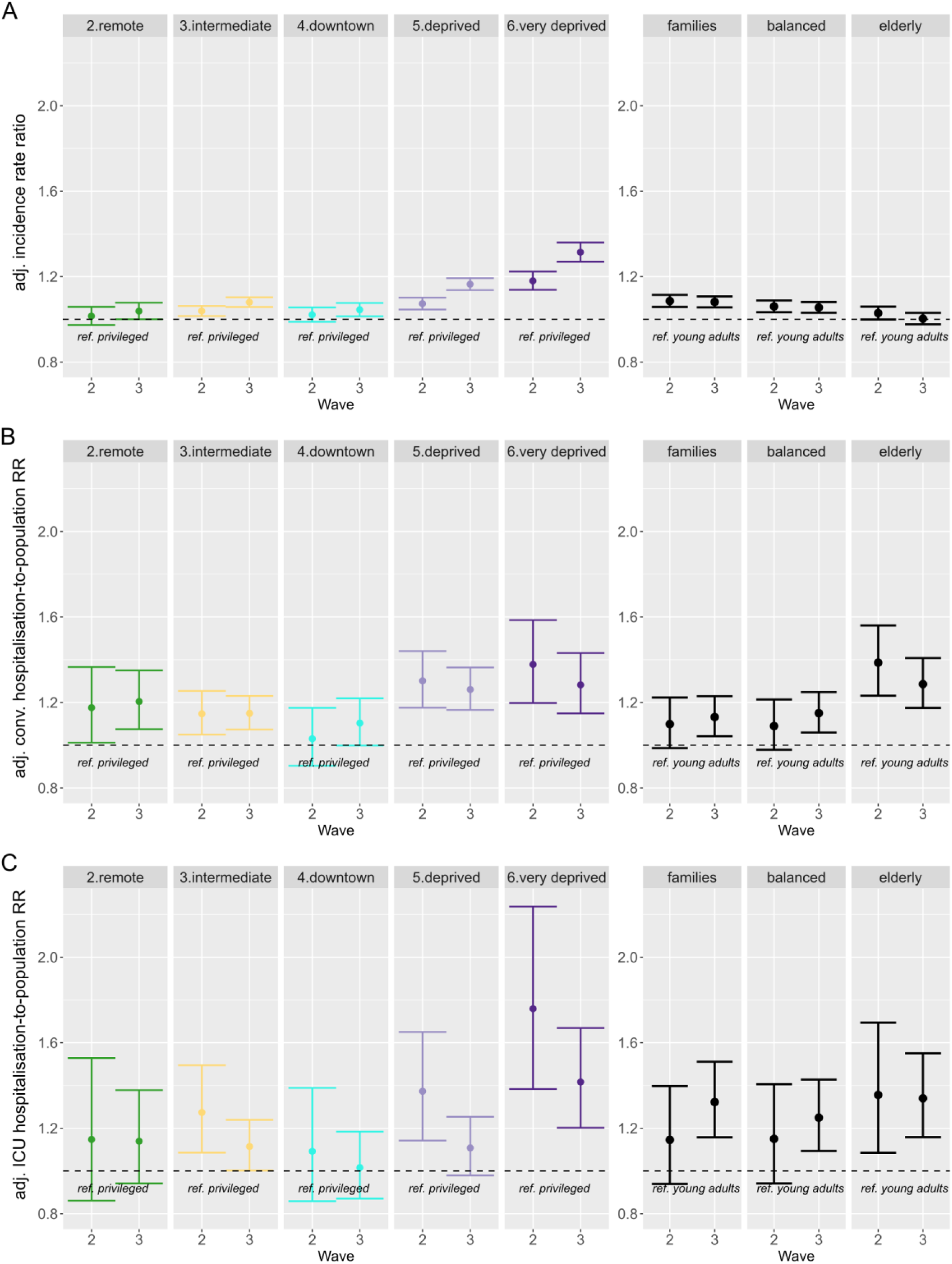
Forest plots of (A) adjusted incidence rate ratio, (B) adjusted conventional hospitalisation to population rate ratio, and (C) adjusted ICU hospitalisation to population rate ratio across waves 2 and 3. Left panels show variations according to socio-demographic profiles (reference category, “privileged” profile), while right panels show variations according to age profiles (reference category, “young adults” profile).

Deprived and, to a lesser extent, intermediate IRIS also exhibited significantly higher IRR (1.30 [1.18-1.44]; 1.15 [1.05-1.25]), and conventional HRR (1.26 [1.17-1.36]; 1.15 [1.07-1.23])). In these deprived and intermediate IRIS, elevated ICU HRR were observed during wave 2 but returned to level comparable to privileged IRIS by wave 3 (Fig 3, Table S3-S4).

In remote IRIS, COVID-19 IRR were comparable to privileged ones in wave 2 and slightly higher in wave 3. Conventional HRR was greater in those areas for both waves (wave 2: 1.18 [1.01-1.37]; wave 3: 1.20 [1.07-1.35]), whereas ICU HRR did not differ significantly from privileged IRIS (Fig 3, Table S3-S4).

Regarding age structure, IRR were higher in families and balanced IRIS compared with young adult IRIS. Conventional HRR were markedly elevated in elderly IRIS across both waves (wave 2: 1.39 [1.23-1.56]; wave 3: 1.29 [1.17-1.41]) and moderately higher in families and balanced IRIS during wave 3. ICU HRR were significantly higher in elderly IRIS in both waves (1.36 [1.08-1.69]; 1.34 [1.16-1.55]). Families and balanced IRIS also showed a significant increase in ICU HRR during the third wave (Fig3, Tables S2-S4).

Accessibility to general practitioner, laboratory presence and the number of frontline caregivers did not significantly influence COVID-19 IRR, or HRR in either conventional or ICU settings (Tables S2-S4). Distance to the nearest emergency department was significantly associated with a slight reduction in COVID 19 IRR, conventional HRR, and ICU HRR. Although coefficient was close to one, the cumulative effect could be substantial in areas located several tens of kilometres from emergency services (Tables S2-S4). The presence of retirement home was associated with higher conventional HRR but not with increased ICU HRR (Tables S3-S4).

Spatial analysis explicitly accounted for autocorrelation between neighbouring IRIS, as evidenced by the non-significant Moran’s I values on model residuals (Tables S9).

### Factors associated with COVID-19 test positivity and hospitalised-case ratios

Proportion of positive test was modelled as a sensitivity analysis. It closely mirrored incidence rate ratios (Fig S5A, Table S5).

To distinguish between increased viral circulation and greater disease severity, we further modelled the proportion of hospitalised patients among confirmed cases, using the proportion-of-hospitalised-cases ratios (pHCR) (Fig S5B,C, Table S6-S7). This analysis confirmed higher pHCR for very deprived IRIS in both conventional and ICU settings. Intermediate and deprived IRIS also showed elevated conventional pHCR in both waves and higher ICU pHCR during wave 2. All these elevated pHCR were slightly lower than HRR, reflecting the contribution of higher incidence in intermediate, deprived and very deprived IRIS. Remote IRIS likewise exhibited higher conventional pHCR in both waves while ICU pHCR remained comparable to privileged IRIS.

Regarding age structure, pHCR decreased slightly overall after adjustment. For conventional hospitalisation, pHCR were no longer elevated for families and balanced IRIS, whereas they remained significant for elderly IRIS in both waves. In ICU settings, pHCR remained elevated for families and elderly IRIS during wave 3.

## Discussion

This geo-epidemiological study analysed COVID 19 incidence and hospitalisation in both conventional and ICU settings at the finest scale available for 2nd and 3rd epidemic waves in the south-eastern French region.

The effects of the intensity of each epidemic wave (reflected by the number of incident cases) and its severity (measured by the proportion of severe cases relative to total cases) could not be fully disentangled. We therefore based our primary analysis on the hospitalisation rate in the general population which captures the overall hospital burden of COVID-19 for each area. This measure, however, combines both incidence and severity components, as increases in either can lead to higher hospitalisation rates. Complementary analysis modelling the proportion of hospitalisation among positive cases, and adjusted for testing rates, were performed to better isolate the contribution of disease severity.

### Socio-demographic determinants

This study highlights clear inequities in COVID-19 outcomes across socio-demographic profiles. Relative to privileged IRIS, intermediate, deprived and very deprived IRIS experienced both higher viral spread and increased severity, as evidenced by elevated incidence and hospitalisation rates in both conventional and ICU settings. By contrast, remote IRIS exhibited only a limited increase in incidence in wave 3, but an excess of conventional hospitalisations in both waves, without evidence of higher ICU hospitalisations. Downtown IRIS displayed patterns largely comparable to privileged areas.

Deprived and very deprived IRIS correspond to urban, densely populated areas, with a high European Deprivation Index (EDI), particularly pronounced in the very deprived IRIS. In contrast, intermediate IRIS are peri-urban, less densely populated and characterized by a lower EDI (18). A progressive gradient was observed in hospitalisation-to-population rate ratios (HRR), increasing from approximately 10% in intermediate to nearly 30% in very deprived IRIS, reinforces the robustness of social deprivation as a key determinant of COVID-19 burden. Importantly, this gradient persisted when looking at the proportion-of-hospitalised-cases ratios (pHCR), suggesting that the excess was not solely attributable to higher incidence but also reflected structurally greater severity.

These findings are consistent with prior reported evidence, particularly in American and British settings (7,8,35). In the European settings, research conducted in Switzerland and recent longitudinal data from Sweden have similarly demonstrated that residing in socioeconomically disadvantaged areas is associated with increased COVID-19 severity (36,37), although these analysis did not account for spatial autocorrelation. Additionally, a recent nationwide French study by Smaïli et al. examined the relationship between COVID-19 outcome and social deprivation at the IRIS scale (Nature Communication Medicine, accepted). Their findings similarly reported a clear socioeconomic gradient with higher risk of COVID 19 hospitalisation and ICU admission in the most deprived area, using a Bayesian modelling approach. Several methodological differences, however, can be noted. First, areas were classified solely according to social deprivation, using EDI quintiles. This approach notably omitted population density, a factor that could potentially lead to misclassification between urban and rural contexts. By contrast, our HCPC profile classification likely provided more contextually appropriate characterization of area types within the PACA region. Specifically, the “remote” profile captures characteristics of certain rural area that socioeconomic data alone may not fully reflect.

Second, while adjustments were made on vaccination coverage and age distribution, no adjustments were made for healthcare access or the presence of retirement homes. Taken together, these converging results, despite methodological differences, reinforce the robustness of the association between area-level deprivation and COVID-19 outcomes.

Several mechanisms may underly this association. Deprived and very deprived profiles included a higher proportion of blue-collar workers engaged in essential occupations, limiting opportunities for remote work, and social distancing, as well as higher rates of overcrowded housing (18). These patterns, consistent with findings from other international studies (38,39), likely facilitated greater viral transmission. Moreover in deprived populations, multiple and complex barriers – including discrimination, low health literacy, language and cultural obstacles, as well as competing needs and priorities (40–42) – may reduce effective healthcare utilisation. Such barriers may also help explain the delayed decline in hospitalisations observed during lockdowns, particularly the third one, when deprived and very deprived IRIS continued to experience rising admission rates, whereas other IRIS showed a rapid decline. Difficulties in adhering to restrictive measures, shaped by socioeconomic and political context, may have sustained higher transmission and severity in these areas. Additionally, these population carry a higher burden of chronic comorbidities, such as diabetes, obesity and chronic pulmonary disease (43–45) which are well established determinant of COVID-19 severity (46,47). Collectively, these factors likely contribute to the excess severity observed in the most deprived IRIS.

Interestingly, this excess severity appeared markedly lower during the third epidemic wave compared with the second. Early 2021, regional health authorities implemented targeted COVID-19 mediation programs in epidemic hotspots, where mediators carried out awareness campaign, contact-tracing and screening activities (48,49). This could partly explain, in addition to the COVID-19 global improvement of knowledge, the narrowing of the gap. Additionally, a harvesting effect may have reduced ICU admissions in deprived areas, as high-risk individuals affected during wave 2 would have acquired immunity by wave 3. Differential immunisation by socioeconomic status previously documented by the French EpiCoV study between wave 1 and 2 (50), may have similarly occurred in the PACA region between wave 2 and 3.

Remote IRIS are characterized by very low population density, a high proportion of agricultural workers and retirees, relatively high median income and limited access to healthcare services (18). After adjustment on testing rate, age structures and healthcare access, COVID-19 incidence in these areas showed only a modest increase during the third wave compared with privileged IRIS. By contrast, conventional HRR were markedly elevated, and this increase remained stable across both epidemic waves, without a corresponding rise in ICU HRR. Here also, the same variations are found when looking at pHCR. This unexpected isolated increase in conventional hospitalisation rate ratios may reflect differences in clinical decision-making for remote populations, where hospitalisation threshold could be lower due to geographical remoteness. This interpretation is further supported by the absence of ICU HRR increase, and by the specific temporal dynamics of hospitalisation rates, which – unlike those in deprived and very deprived IRIS – declined rapidly during the third lockdown. These findings are consistent with results currently on preparation, which focus specifically on rural populations (Schalwijk, Gaudart et al., report on preparation)

### Age structure

Regarding age structure, the higher HRR observed in “elderly” IRIS are in line with previous evidence, as advanced age is a well-established determinant of COVID-19 severity (46,51). Variation in hospitalisation for families and balanced IRIS, seem to be partially depending on incidence rate, as significant higher rates in conventional HRR for both waves are not observed in conventional pHCR for those IRIS.

### Adjusting variables

The presence of a retirement home within an IRIS was consistently associated with an approximately 10% increase in conventional hospital admission during both waves, but no increase in ICU admission. This apparent paradox could be explained by a lower rate of ICU admissions among very elderly and fragile individuals (51,52). In this context, the distinction between legitimate refusals of non-beneficial intensive care, and triage decisions imposed by limited ICU capacity is difficult to establish.

### Study limitation

This study has several limitations. First, certain hidden population groups were not accounted for, as they were absent from the available census datasets. This well-recognised intrinsic bias is inherent to all ecological studies (53). Recent studies focusing on COVID-19 circulation among homeless population in Marseille and in the French territory may, however, help to better characterise this specific hidden group (54,55). Moreover, IRIS units were not always homogeneous; for instance, some downtown districts of Marseille include informal settlements or slum areas that may not be fully represented by aggregated socio-economic indicators. Because addresses were self-reported and geocoded automatically, some degree of misclassification cannot be excluded, although the large population base likely limited its influence on the results.

Additionally, although conventional hospitalisation is primarily determined by clinical criteria, the potential influence of socio-demographic factors on admission decisions cannot be fully disentangled in this study. Socially deprived or isolated patients may be more frequently hospitalised in conventional settings to ensure adequate treatment and monitoring. However, the elevated ICU hospitalisation rates observed in the most deprived IRIS suggest a true severity component, which was not evident in remote areas. Further research is warranted to better understand healthcare pathways and the factors influencing medical decision-making in both deprived and rural contexts.

The healthcare accessibility indicators used for model adjustment (LPA, density of primary healthcare professional, average distance to emergency services) do not seem to play a major role. Yet, these variables capture only healthcare availability and not its effective use. As noted above, barriers to care are particularly relevant in deprived populations, and the absence of an observed effect may mask underlying disparities in actual healthcare use.

Information on vaccination coverage at a fine spatial resolution was also lacking. Vaccination campaign began in early 2021, coinciding with the onset of the third wave, initially targeting the most vulnerable populations in terms of age and comorbidities, and uptake was generally higher in wealthier groups (56,57). Although this could have influenced results, the observed narrowing of disparities in severity between the second and third waves indicates that any confounding effect was likely limited.

Information on ethnicity is not collected in France. However, the socio-demographic profiles incorporated information on the proportion of immigrant and foreigners, who are overrepresented in deprived and very deprived IRIS. Given that minority ethnic group in France, particularly residents of African origin, have been shown to face increased risks of severe COVID-19 and mortality (11,58), part of this effect may also have been indirectly captured through socioeconomic categorisation.

The two IRIS profile variables – sociodemographic level and age structure – were colinear, raising the possibility of coefficient instability. Indeed, more than one third of IRIS classified as “familial” also belonged to either “deprived” or “very deprived” profiles. This may explain, for example, the apparent isolated increase in ICU pHCR in “familial” IRIS during the third wave, which coincided with a decrease in “deprived” and “very deprived” IRIS. Nevertheless, the consistency of the findings across several analytical models suggests that collinearity had only a limited impact on the robustness of the estimated coefficient.

## Conclusion

In conclusion, this fine-scale ecological study provides an integrative understanding of COVID-19 incidence and severity across the PACA region. The HCPC-based classification enabled the identification of areas sharing similar demographic and socio-economic characteristics, offering a relevant framework for regional public health action. A clear sociodemographic gradient was observed, with increasing viral spread and disease severity from intermediate to deprived and very deprived areas, while privileged and central urban areas were less affected. Remote areas showed elevated conventional hospitalisation rates that may reflect clinical decisions influenced by geographic remoteness rather than increased severity. These findings, consistent with previous studies, emphasize the need for tailored prevention strategies in socially vulnerable areas and call for preparedness measures to better address health inequalities in future pandemics. In the PACA region, targeted interventions should particularly focus on very deprived and deprived IRIS, which, despite representing only around 7.1% and 18% of the population (and 0.20 % and 4.1% of the region’s area respectively), bear a disproportionate burden of severe disease.

## Ethical statement

To ensure the confidentiality of the information and in accordance with French regulations, only data aggregated to the IRIS and to the day were processed for this study. Clearance was obtained through a specific convention (number 22DIRA41-0) between Aix Marseille University and Santé publique France, from the Aix Marseille University Ethic committee (number 2022-10-20-006), and from the Aix Marseille University Data Protection Officer (number 513087).

## Data Availability

All data produced in the present study are available upon reasonable request to the authors

## Acknowledgments

We aknowledge *Santé publique France*, and the *Agence régionale de santé Provence-Alpes-Côte d’Azur (PACA)* for granting access to the data used in this analysis. We are also deeply greatful to the *Ligue Nationale Contre le Cancer*, for providing the European Deprivation Index (EDI). Our sincere thanks go to F. Danjou, A. Ramdani, and P. Malfait for their assistance in data processing and preparation.

## Authors’ contributions

JG designed the study and supervised its implementation. SN, AR, MH, FD and FF prepared the SI-DEP and SI-VIC datasets at the IRIS level. GG and PG performed the statistical analyses and drafted the manuscript, with contributions from JL and JG, with the help of LC and PS. All authors participated the interpretations of the results, reviewed and approved the final version of the manuscript.

