## supplementary data for "Geoepidemiology of COVID-19 hospitalisations and severity: impact of social deprivation and remote areas, in the south-eastern Region, France"

**Table S1**: Database sources

| Database name | Date of publication | Last access to the database | Author | URL |
|---|---|---|---|---|
| **Equipment** | 12/07/2021 | 13/07/2021 | INSEE | https://www.insee.fr/fr/statistiques/3568629?sommaire=3568656&q=bpe+2020 |
| **Census** | 09/12/2020 | 12/04/2021 | INSEE | https://www.insee.fr/fr/statistiques/4515565?sommaire=4516122&q=recensement+2017#consulter |
| **Housing** | 09/12/2020 | 21/05/2021 | INSEE | https://www.insee.fr/fr/statistiques/4515532?sommaire=4516107&q=base+logement#dictionnaire |
| **European deprivation index (EDI)** | 2015 | | MapInMed | https://anticipe.eu/plateformes/MAPinMED |
| **Vaccination** | 10 /09/2021 | 20/09/2021 | | https://datavaccin-covid.ameli.fr/explore/?exclude.theme=Datavisualisation&sort=modified |
| **IRIS borders (shapefile)** | | 21/05/2021 | French Government | https://www.data.gouv.fr/en/datasets/decoupage-iris-combine-aux-limites-communales-openstreetmap/ |
| **Localised Potential Access (LPA)** | 02/03/2020 | 30/07/2021 | DREES | https://drees2-sgsocialgouv.opendatasoft.com/explore/dataset/530_l-accessibilite-potentielle-localisee-apl/information/ |

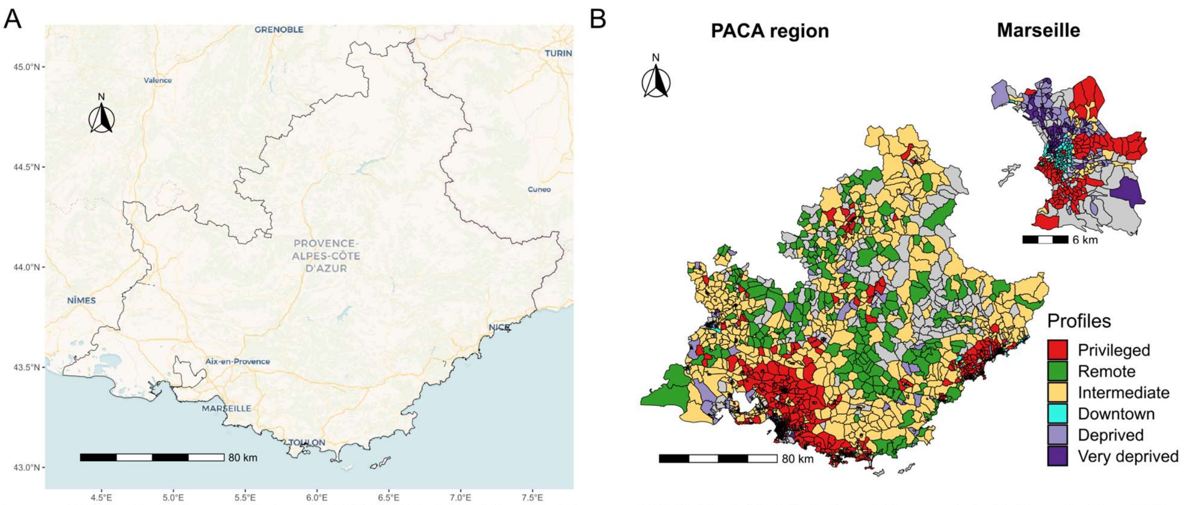


Source : © OpenStreetMap contributors, © CARTO (CC BY 4.0) license, data.gouv, INSEE, MapInMed / Author : P Garneret / Edition : October 2025

**Figure. S1**: Geographic overview of the Provence-Alpes-Côte d'Azur (PACA) region. (A) Map of the PACA region and its main cities. (B) Spatial distribution of socio-demographic profiles across the PACA region, with a detailed focus on Marseille. Grey areas correspond to IRIS unit with no hospitalisation data reported.

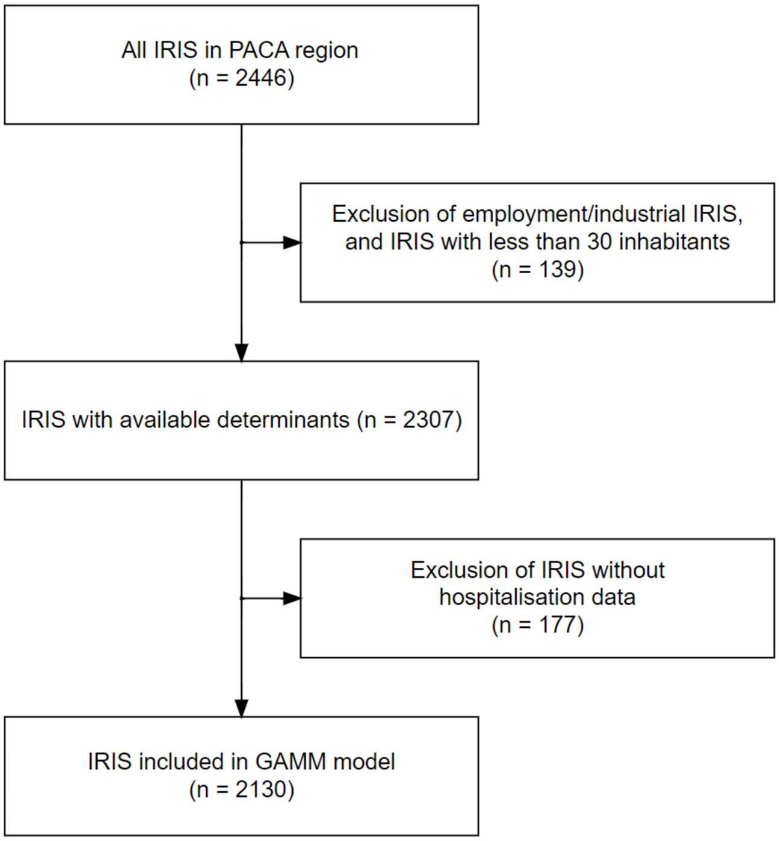


**Figure S2:** Flow chart of IRIS selection.
2130 IRIS (4.92 million inhabitants) were included among the 2446 IRIS in PACA region (5.04 million inhabitants).

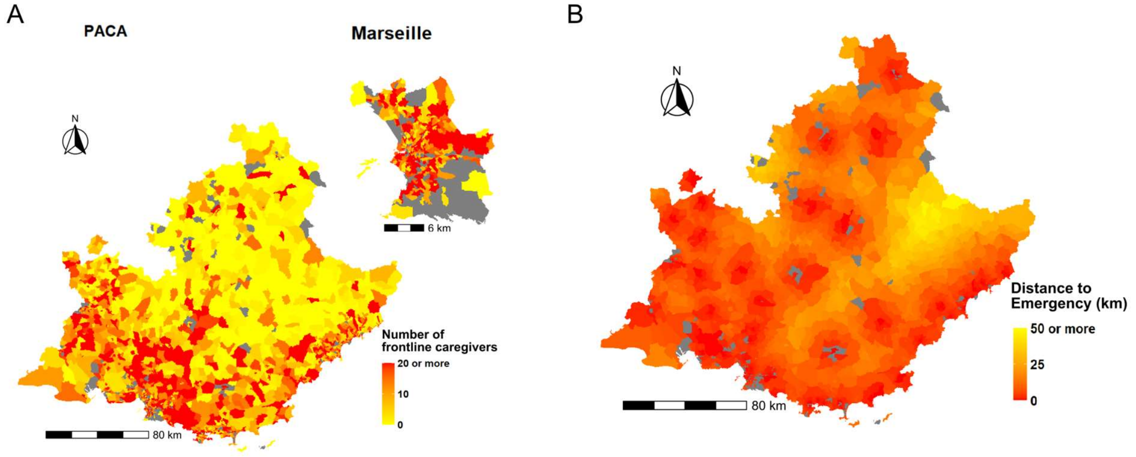


Source : data.gouv, INSEE / Author : G Gaubert / Edition : October 2025

**Figure S3 :** (A) Map of the number of frontline caregivers per IRIS, in PACA region and its main city Marseille. The NA (grey) class correspond to unpopulated areas (B) Map of the distance to Emergency reception service for each service, at the IRIS level in PACA region. The NA (grey) class correspond to unpopulated areas. Distances were calculated between IRIS centroids.

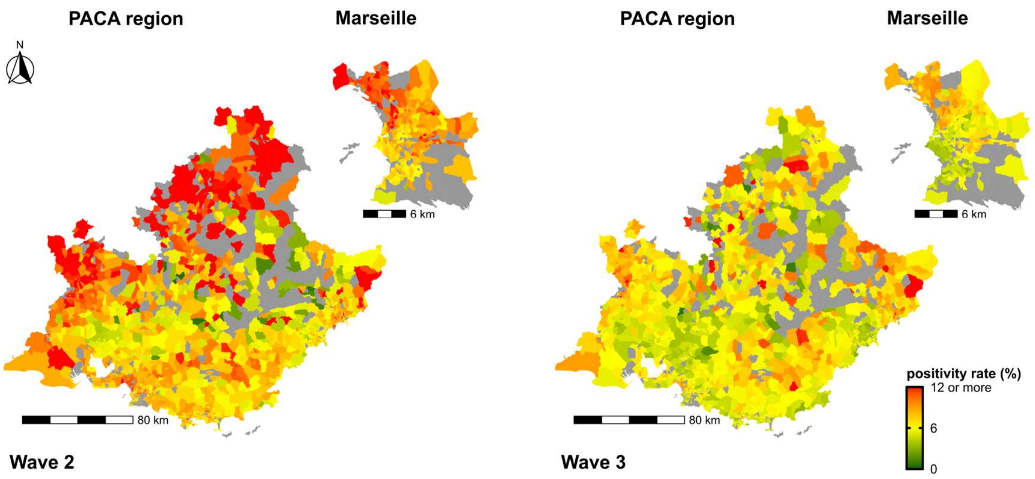


Source : data.gouv, SI-DEP / Author : P Garneret / Edition : October 2025

**Figure S4 :** Spatial distribution of COVID-19 positivity rate, in the PACA region, with a focus on Marseille, during epidemic waves 2 and 3. Grey areas correspond to IRIS unit with no hospitalisation data reported.

**Table S2:** Adjusted incidence rate ratios (IRR) for COVID-19 infection

| | | wave 2 | | | wave 3 | | |
|---|---|---|---|---|---|---|---|
| **Variable** | **Modality** | **SIR** | **CI95** | **p** | **SIR** | **CI95** | **p** |
| Socioeconomic profile (ref : Privileged) | Remote | 1.01 | 0.97 - 1.06 | 0.489 | 1.04 | 1 - 1.08 | 0.048 |
| | Intermediate | 1.04 | 1.02 - 1.06 | 0.001 | 1.08 | 1.06 - 1.1 | < 0.001 |
| | Downtown | 1.02 | 0.99 - 1.06 | 0.208 | 1.04 | 1.01 - 1.08 | 0.004 |
| | Deprived | 1.07 | 1.05 - 1.1 | < 0.001 | 1.16 | 1.14 - 1.19 | < 0.001 |
| | Very deprived | 1.18 | 1.14 - 1.22 | < 0.001 | 1.31 | 1.27 - 1.36 | < 0.001 |
| Age structure profile (ref : Young adults) | Families | 1.09 | 1.06 - 1.11 | < 0.001 | 1.08 | 1.06 - 1.11 | < 0.001 |
| | Balanced | 1.06 | 1.03 - 1.09 | < 0.001 | 1.06 | 1.03 - 1.08 | < 0.001 |
| | Elderly | 1.03 | 1 - 1.06 | 0.056 | 1.00 | 0.98 - 1.03 | 0.82 |
| Accessibility to general practitioner | | 1.01 | 1 - 1.02 | 0.087 | 1.00 | 0.99 - 1.01 | 0.968 |
| Retirement home presence | | 0.98 | 0.97 - 1 | 0.018 | 0.98 | 0.96 - 0.99 | 0.001 |
| Distance to Emergency reception (km) | | 0.99 | 0.99 - 1 | 0.004 | 1.00 | 0.99 - 1 | 0.08 |
| Laboratory presence | | 1.02 | 1 - 1.04 | 0.028 | 1.00 | 0.99 - 1.02 | 0.911 |
| Number of frontline caregivers | | 1.00 | 1 - 1 | 0.032 | 1.00 | 1 - 1 | 0.008 |

**Table S3:** Adjusted hospitalisation-to population rate ratio (HRR) for COVID-19 in conventional settings.

| | | wave 2 | | | wave 3 | | |
|---|---|---|---|---|---|---|---|
| **Variable** | **Modality** | **SIR** | **CI95** | **p** | **SIR** | **CI95** | **p** |
| Socioeconomic profile (ref : Privileged) | Remote | 1.18 | 1.01 - 1.37 | 0.035 | 1.20 | 1.07 - 1.35 | 0.001 |
| | Intermediate | 1.15 | 1.05 - 1.25 | 0.003 | 1.15 | 1.07 - 1.23 | < 0.001 |
| | Downtown | 1.03 | 0.9 - 1.17 | 0.656 | 1.10 | 1 - 1.22 | 0.054 |
| | Deprived | 1.30 | 1.18 - 1.44 | < 0.001 | 1.26 | 1.17 - 1.36 | < 0.001 |
| | Very deprived | 1.38 | 1.2 - 1.59 | < 0.001 | 1.28 | 1.15 - 1.43 | < 0.001 |
| Age structure profile (ref : Young adults) | Families | 1.10 | 0.99 - 1.22 | 0.087 | 1.13 | 1.04 - 1.23 | 0.003 |
| | Balanced | 1.09 | 0.98 - 1.21 | 0.119 | 1.15 | 1.06 - 1.25 | 0.001 |
| | Elderly | 1.39 | 1.23 - 1.56 | < 0.001 | 1.29 | 1.17 - 1.41 | < 0.001 |
| Accessibility to general practitioner | | 1.00 | 0.96 - 1.03 | 0.83 | 1.01 | 0.99 - 1.04 | 0.356 |
| Retirement home presence | | 1.16 | 1.09 - 1.23 | < 0.001 | 1.08 | 1.03 - 1.13 | 0.001 |
| Distance to Emergency reception (km) | | 0.98 | 0.98 - 0.99 | < 0.001 | 0.99 | 0.99 - 1 | 0.029 |
| Laboratory presence | | 1.02 | 0.95 - 1.09 | 0.556 | 1.03 | 0.98 - 1.08 | 0.305 |
| Number of frontline caregivers | | 1.00 | 1 - 1 | 0.137 | 1.00 | 1 - 1 | 0.056 |

**Table S4:** Adjusted hospitalisation-to population rate ratio (HRR) for COVID-19 in ICU settings

| | | wave 2 | | | wave 3 | | |
|---|---|---|---|---|---|---|---|
| **Variable** | **Modality** | **SIR** | **CI95** | **p** | **SIR** | **CI95** | **p** |
| Socioeconomic profile (ref : Privileged) | Remote | 1.15 | 0.86 - 1.53 | 0.347 | 1.14 | 0.94 - 1.38 | 0.18 |
| | Intermediate | 1.27 | 1.09 - 1.49 | 0.003 | 1.11 | 1 - 1.24 | 0.045 |
| | Downtown | 1.09 | 0.86 - 1.39 | 0.474 | 1.02 | 0.87 - 1.18 | 0.845 |
| | Deprived | 1.37 | 1.14 - 1.65 | 0.001 | 1.11 | 0.98 - 1.25 | 0.104 |
| | Very deprived | 1.76 | 1.38 - 2.24 | < 0.001 | 1.42 | 1.2 - 1.67 | < 0.001 |
| Age structure profile (ref : Young adults) | Families | 1.15 | 0.94 - 1.4 | 0.18 | 1.32 | 1.16 - 1.51 | < 0.001 |
| | Balanced | 1.15 | 0.94 - 1.41 | 0.17 | 1.25 | 1.09 - 1.43 | 0.001 |
| | Elderly | 1.36 | 1.08 - 1.69 | 0.007 | 1.34 | 1.16 - 1.55 | < 0.001 |
| Accessibility to general practitioner | | 1.00 | 0.95 - 1.06 | 0.883 | 1.03 | 0.99 - 1.07 | 0.123 |
| Retirement home presence | | 0.95 | 0.85 - 1.07 | 0.391 | 1.00 | 0.93 - 1.08 | 0.98 |
| Distance to Emergency reception (km) | | 1.00 | 0.99 - 1.01 | 0.929 | 0.99 | 0.97 - 1 | 0.008 |
| Laboratory presence | | 1.00 | 0.88 - 1.13 | 0.957 | 1.02 | 0.94 - 1.11 | 0.606 |
| Number of frontline caregivers | | 1.00 | 1 - 1.01 | 0.151 | 1.00 | 1 - 1 | 0.976 |

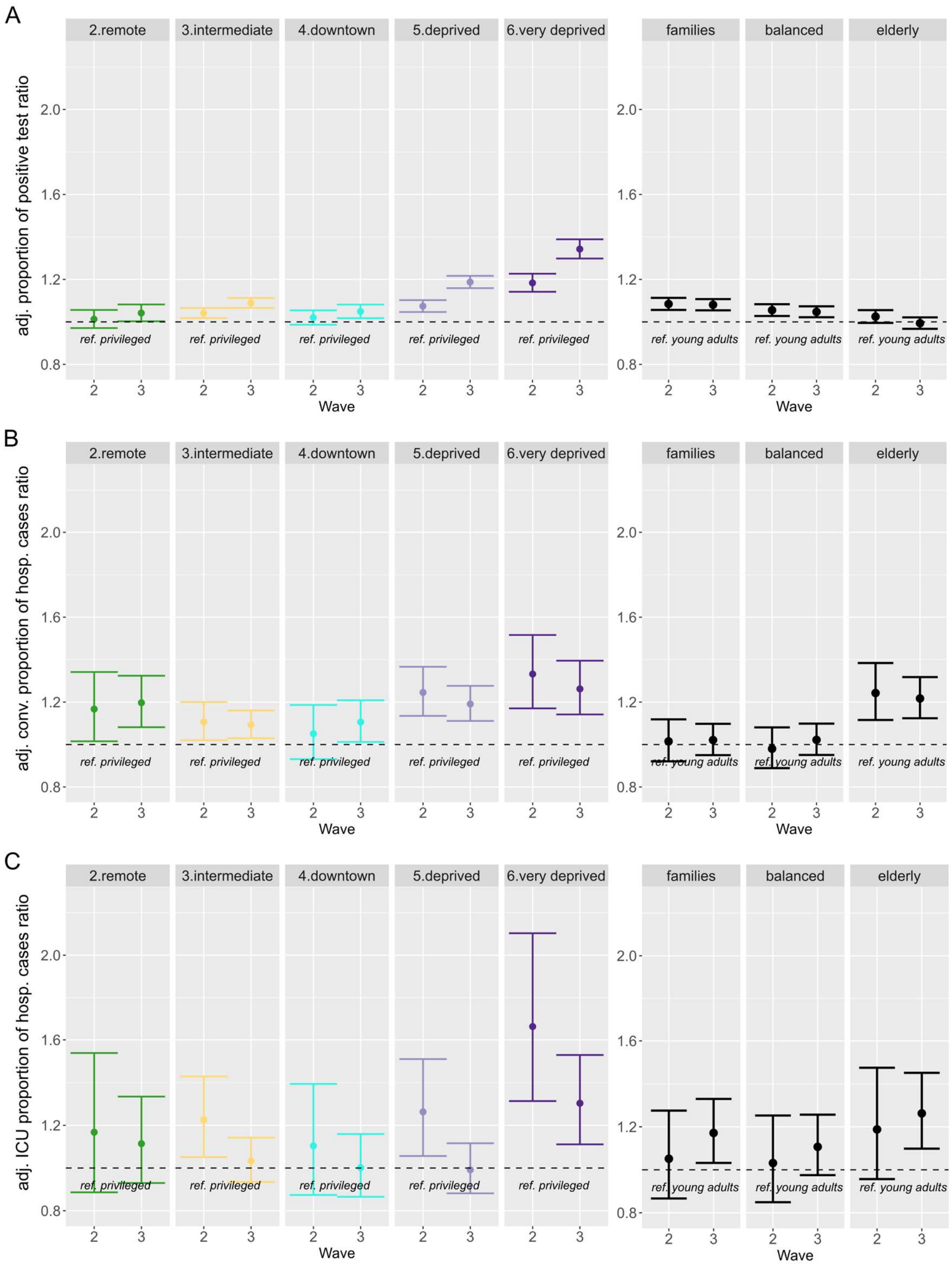


**Figure S5:** Forest plots of (A) adjusted proportion of positive test ratio, (B) adjusted proportion of hospitalised cases ratio in conventional settings, and (C) adjusted proportion of hospitalised cases ratio in ICU settings, across waves 2 and 3. Left panels show variations according to sociodemographic profiles (reference category, “privileged” profile), while right panels show variations according to age profiles (reference category, “young adults” profile).

**Table S5:** Adjusted proportion of positive tests ratio for COVID-19

| | | wave 2 | | | wave 3 | | |
|---|---|---|---|---|---|---|---|
| **Variable** | **Modality** | **SIR** | **CI95** | **p** | **SIR** | **CI95** | **p** |
| Socioeconomic profile (ref : Privileged) | Remote | 1.01 | 0.97 - 1.06 | 0.557 | 1.04 | 1 - 1.08 | 0.035 |
| | Intermediate | 1.04 | 1.02 - 1.07 | 0.001 | 1.09 | 1.07 - 1.11 | < 0.001 |
| | Downtown | 1.02 | 0.99 - 1.05 | 0.245 | 1.05 | 1.02 - 1.08 | 0.002 |
| | Deprived | 1.07 | 1.05 - 1.1 | < 0.001 | 1.19 | 1.16 - 1.22 | < 0.001 |
| | Very deprived | 1.18 | 1.14 - 1.23 | < 0.001 | 1.34 | 1.3 - 1.39 | < 0.001 |
| Age structure profile (ref : Young adults) | Families | 1.08 | 1.06 - 1.11 | < 0.001 | 1.08 | 1.05 - 1.11 | < 0.001 |
| | Balanced | 1.06 | 1.03 - 1.08 | < 0.001 | 1.05 | 1.02 - 1.07 | < 0.001 |
| | Elderly | 1.02 | 1 - 1.06 | 0.1 | 0.99 | 0.97 - 1.02 | 0.662 |
| Accessibility to general practitioner | | 1.01 | 1 - 1.02 | 0.07 | 1.00 | 0.99 - 1.01 | 0.852 |
| Retirement home presence | | 0.98 | 0.96 - 0.99 | 0.005 | 0.97 | 0.96 - 0.99 | < 0.001 |
| Distance to Emergency reception (km) | | 0.99 | 0.99 - 1 | 0.002 | 1.00 | 0.99 - 1 | 0.094 |
| Laboratory presence | | 1.02 | 1 - 1.03 | 0.034 | 1.00 | 0.98 - 1.02 | 0.962 |
| Number of frontline caregivers | | 1.00 | 1 - 1 | 0.033 | 1.00 | 1 - 1 | 0.003 |

**Table S6:** Adjusted proportion of hospitalised cases ratio (pHCR) for COVID-19 in conventional settings

| | | wave 2 | | | wave 3 | | |
|---|---|---|---|---|---|---|---|
| **Variable** | **Modality** | **SIR** | **CI95** | **p** | **SIR** | **CI95** | **p** |
| Socioeconomic profile (ref : Privileged) | Remote | 1.17 | 1.02 - 1.34 | 0.03 | 1.20 | 1.08 - 1.32 | 0.001 |
| | Intermediate | 1.11 | 1.02 - 1.2 | 0.015 | 1.09 | 1.03 - 1.16 | 0.004 |
| | Downtown | 1.05 | 0.93 - 1.19 | 0.42 | 1.11 | 1.01 - 1.21 | 0.026 |
| | Deprived | 1.25 | 1.14 - 1.37 | < 0.001 | 1.19 | 1.11 - 1.28 | < 0.001 |
| | Very deprived | 1.33 | 1.17 - 1.52 | < 0.001 | 1.26 | 1.14 - 1.39 | < 0.001 |
| Age structure profile (ref : Young adults) | Families | 1.01 | 0.92 - 1.12 | 0.764 | 1.02 | 0.95 - 1.1 | 0.569 |
| | Balanced | 0.98 | 0.89 - 1.08 | 0.687 | 1.02 | 0.95 - 1.1 | 0.553 |
| | Elderly | 1.24 | 1.12 - 1.38 | < 0.001 | 1.22 | 1.12 - 1.32 | < 0.001 |
| Accessibility to general practitioner | | 0.98 | 0.95 - 1.02 | 0.319 | 1.01 | 0.98 - 1.03 | 0.677 |
| Retirement home presence | | 1.10 | 1.04 - 1.16 | 0.001 | 1.08 | 1.04 - 1.13 | < 0.001 |
| Distance to Emergency reception (km) | | 0.99 | 0.98 - 1 | 0.093 | 1.00 | 0.99 - 1 | 0.318 |
| Laboratory presence | | 1.00 | 0.94 - 1.06 | 0.874 | 1.00 | 0.96 - 1.05 | 1 |
| Number of frontline caregivers | | 1.00 | 1 - 1 | 0.302 | 1.00 | 1 - 1 | 0.037 |

**Table S7:** Adjusted proportion of hospitalised cases ratio (pHCR) for COVID-19 in ICU settings

| | | wave 2 | | | wave 3 | | |
|---|---|---|---|---|---|---|---|
| **Variable** | **Modality** | **SIR** | **CI95** | **p** | **SIR** | **CI95** | **p** |
| Socioeconomic profile (ref : Privileged) | Remote | 1.17 | 0.89 - 1.54 | 0.271 | 1.11 | 0.93 - 1.33 | 0.242 |
| | Intermediate | 1.23 | 1.05 - 1.43 | 0.009 | 1.03 | 0.93 - 1.14 | 0.523 |
| | Downtown | 1.10 | 0.87 - 1.39 | 0.408 | 1.00 | 0.87 - 1.16 | 0.983 |
| | Deprived | 1.26 | 1.06 - 1.51 | 0.011 | 0.99 | 0.88 - 1.12 | 0.89 |
| | Very deprived | 1.66 | 1.31 - 2.1 | < 0.001 | 1.30 | 1.11 - 1.53 | 0.001 |
| Age structure profile (ref : Young adults) | Families | 1.05 | 0.87 - 1.28 | 0.611 | 1.17 | 1.03 - 1.33 | 0.014 |
| | Balanced | 1.03 | 0.85 - 1.25 | 0.755 | 1.11 | 0.98 - 1.26 | 0.116 |
| | Elderly | 1.19 | 0.96 - 1.47 | 0.119 | 1.26 | 1.1 - 1.45 | 0.001 |
| Accessibility to general practitioner | | 1.00 | 0.95 - 1.05 | 0.904 | 1.03 | 0.99 - 1.07 | 0.144 |
| Retirement home presence | | 0.90 | 0.8 - 1 | 0.059 | 1.00 | 0.93 - 1.08 | 0.967 |
| Distance to Emergency reception (km) | | 1.01 | 1 - 1.02 | 0.149 | 0.99 | 0.98 - 1 | 0.097 |
| Laboratory presence | | 0.98 | 0.87 - 1.11 | 0.743 | 1.00 | 0.93 - 1.09 | 0.908 |
| Number of frontline caregivers | | 1.00 | 1 - 1 | 0.316 | 1.00 | 1 - 1 | 0.913 |

**Table S8:** Moran's I estimates of spatial autocorrelation for COVID-19 indicators across IRIS in the PACA region, stratified by epidemic wave

| Indicator | Wave | Moran's Index | p-value |
|---|---|---|---|
| Incidence rates | 2 | 0,29 | < 0.001 |
| | 3 | 0,41 | < 0.001 |
| Conventional hospitalisation-to-population rate | 2 | 0,16 | < 0.001 |
| | 3 | 0,08 | < 0.001 |
| ICU hospitalisation-to-population rate | 2 | 0 | 0.477 |
| | 3 | 0,1 | < 0.001 |

**Table S9:** Moran's I estimates of spatial autocorrelation in GAMM residuals for COVID-19 indicators across IRIS in the PACA region, stratified by epidemic wave

| Indicator | Wave | Moran's Index | p-value |
|---|---|---|---|
| Incidence rates | 2 | -0,01 | 0.724 |
| | 3 | 0,04 | 0.004 |
| Conventional hospitalisation-to-population rate | 2 | -0,02 | 0.939 |
| | 3 | -0,04 | 0.996 |
| ICU hospitalisation-to-population rate | 2 | -0,02 | 0.908 |
| | 3 | -0,02 | 0.944 |
